# Motivational processes involved in long-term exercise engagement: A multiple case study testing Self-Determination Theory in a sample of older adults with mild cognitive impairment and early dementia

**DOI:** 10.1101/2025.10.07.25337231

**Authors:** Jennie E Hancox, Claudio Di Lorito, Veronika van der Wardt, Kristian Pollock, Kavita Vedhara, Rowan H Harwood

**Affiliations:** Division of Primary Care, School of Medicine, University of Nottingham, Nottingham, UK; Division of Rehabilitation, Aging and Well-being, School of Medicine, University of Nottingham, Nottingham, UK; School of Health Sciences, University of Nottingham, Nottingham, UK

**Keywords:** motivation, self-determination theory, exercise, dementia, older adults, adherence, physical activity, case study

## Abstract

Exercise and physical activity are thought to confer widespread health benefits such as, slowing cognitive and physical decline and reducing falls in people with mild cognitive impairment (MCI) and early dementia. However, long-term exercise and physical activity engagement in this population is poor. This study aimed to explore the motivational processes involved in long-term engagement to a home-based exercise and activity intervention among people with MCI and early dementia and to test the validity of Self-determination Theory (SDT) propositions. Multiple case studies spanning 24 months were undertaken with a sample of 12 participants and their carers. Qualitative data (face-to-face interviews and telephone calls) and quantitative data (exercise diaries) were collected. Qualitative interviews were transcribed and analysed using thematic analysis. Descriptive statistics were calculated for quantitative data and used to enhance interpretation of case studies. Data analysis was performed using pattern matching logic in which the findings of each case study were compared with the theoretical propositions of SDT. Cross-case synthesis identified four profiles of participants: Long-term engagement - continuation of exercises and physical activities for 24 months; long-term partial engagement - stopped the exercises after 12 months, but continued the physical activities for 24 months; short-term engagement - stopped the exercises and physical activities after 12 months; lack of engagement - exercises and physical activities discontinued by 6 months. SDT could be used to understand the motivational processes involved in long-term engagement. Aligned with SDT, the provision of basic needs support over time and intrinsic motivation were crucial for long-term exercise and physical activity engagement. Implications for researchers, practitioners and policy makers supporting older adults with MCI or dementia to engage in physical activity and/or exercise long-term are discussed.

## Introduction

A physically active lifestyle is associated with a range of health benefits, including improved physical function and psychological well-being and reduced incidence of many chronic conditions (e.g., diabetes, dementia). For people with mild cognitive impairment (MCI; impairment of function in cognition but that does not fully meet the criteria for dementia; Albert et al., 2011) and early dementia (a progressive clinical syndrome characterised by irreversible deterioration in an individual’s cognitive abilities, such as short-term memory and executive function, which interferes with the ability to perform everyday activities; WHO, 2017) physical activity and therapeutic exercise is important for increasing functional ability (Pitkala et al., 2013), cognition (Lautenschlager et al., 2008), mood (Hernández et al., 2015) and reducing falls risk (Sherrington et al., 2011). However, this population has been found to have low activity levels and to experience a significant decline in physical activity and exercise over time (Sabia et al., 2017). Physical activity is broadly defined as ‘any bodily movement produced by skeletal muscles that results in energy expenditure’ (Caspersen, Powell & Christenson, 1985). In this study we focus on leisure-time physical activity which is described by Caspersen et al. (1985) to include sports (e.g., running), household tasks (e.g., gardening), daily activities (e.g., shopping) and conditioning exercises (e.g., leg-raises).

Exercise and physical activity interventions aiming to increase activity levels in older adults with MCI or dementia have ranged in duration from 6-80 weeks (mean = 23 weeks) and have reported varied levels of adherence, ranging from 16-100% (mean = 70%) (Di Lorito et al., under review). A limitation of previous studies is that they have tended to measure adherence to exercise sessions only and do not consider participation in other leisure-time physical activities (e.g., sports, household tasks or daily activities) or assess engagement in exercise once the intervention had finished. Furthermore, previous studies have rarely considered why people do or do not engage in the long-term and the role of motivation in sustaining exercise levels over time.

Theories of motivation and behaviour change can be helpful for understanding the mechanisms underlying engagement. This understanding can be used to design and deliver interventions in a way which is more likely to promote adherence. Self-determination theory (SDT; Ryan & Deci, 2017) is a theory of motivation, which is particularly relevant to understanding long-term adherence in the physical activity domain (Teixeira et al., 2012). Previous cross-sectional research has suggested SDT may be a suitable framework to explore the mechanisms underlying activity engagement in older adults (Kirkland et al., 2011). Self-determination is considered the degree to which a person is the originator of their own behavior and that the behaviour is chosen, initiated and endorsed with an internal locus of causality (Reeve, Nix, & Hamm, 2003).

A key tenet of SDT is that the reasons why an individual may choose to engage in a behavior lie on a continuum of self-determination (Ryan & Deci, 2000; see Figure 1). It is believed that an understanding of these reasons is essential for predicting long-term engagement.

**Figure 1.**
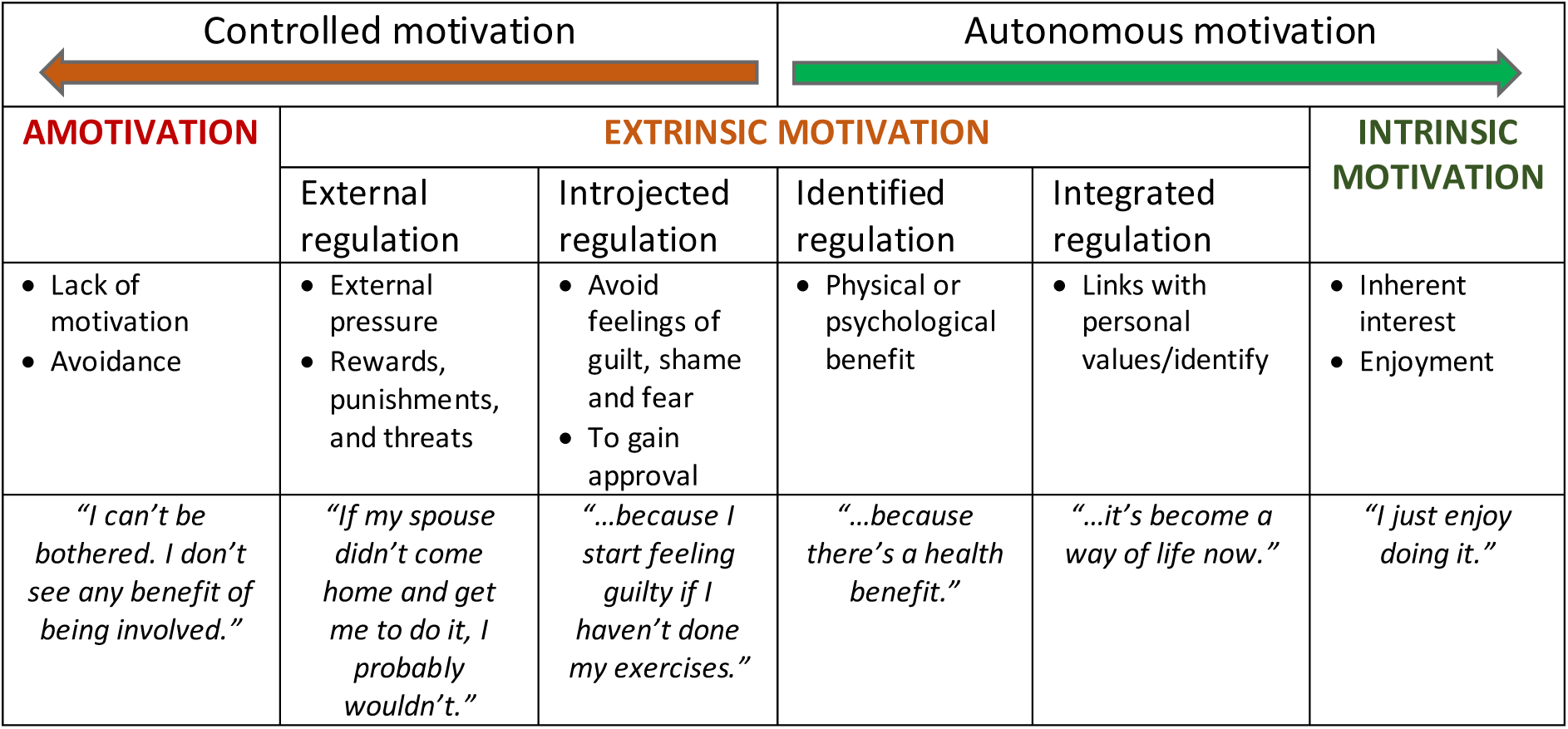
Self-determination theory continuum of motivation regulations (adapted from Ryan & Deci, 2017).

Intrinsic motivation (i.e., engaging for the inherent interest or enjoyment of the activity itself) has been found to be the strongest predictor of long-term physical activity and exercise adherence (Teixeira et al., 2012). The continuum contains four types of extrinsic motivation (see Figure 1). Integrated regulation (because the activity is part of an individuals’ identity and aligned with their interests and values) and identified regulation (because of the associated benefits) are considered to be the more autonomous extrinsic motivations and are associated with psychological well-being (Ng et al., 2012) and continued activity engagement (Rodgers et al., 2010). The more controlled forms of extrinsic motivation are introjected regulation (because of internal pressure, to avoid feelings of guilt or to increase feelings of self-worth) and external regulation (because of external pressure, to please significant others). The relationship between controlled regulations and physical activity and exercise engagement is less clear. Previous research (Kinnafick et al., 2014) has suggested that feelings of guilt for not doing the activity and not wanting to let others down can facilitate adoption of physical activity initially. However, high levels of controlled regulations are not considered conducive to sustained, long-term activity behaviour (Teixeria et al., 2012). Amotivation (a lack of either intrinsic or extrinsic motivation) has been found to be predictive of dropout (Sarrazin, 2002).

Previous cross-sectional research with older adults (Kirkland et al., 2011) found those with more self-determined motivations to report higher levels of physical activity and exercise engagement. However, previous SDT research with older adults has not explored changes in motivational processes over time or the applicability of SDT to older adults with cognitive impairment. Thus, we do not yet know what factors influence the extent to which older adults with MCI and early dementia may become more intrinsically motivated to engage in physical activity and/or exercise long-term and whether SDT is a suitable framework to explore the mechanisms underlying activity persistence in this population.

A sub-theory of SDT, Basic Psychological Needs Theory, posits that intrinsic motivation can be promoted by fulfilling three basic psychological needs:

- Autonomy (feeling a sense of ownership of one’s actions);
- Competence (feeling effective at carrying out actions);
- Relatedness (feeling connected to, cared for and accepted by significant others).

These basic needs are considered to be innate (rather than learned) and universal to all individuals (Ryan & Deci, 2000). The functional significance – the extent to which each of the needs influences individuals’ motivation, however, depends on the specific situation (Ryan & Deci, 2017). For example, Kinnafick et al. (2014) used a longitudinal case study method to explore motivational process involved in the transition of adults from being physically inactive to engaging in a walking programme and found satisfaction of the needs for competence and relatedness to be essential for adoption, but that a sense of autonomy was more pertinent later on in facilitating adults’ long-term adherence to walking.

The extent to which the social-environment supports or thwarts these needs is considered to facilitate or undermine self-determined motivation. For instance, when significant others (e.g., health professional or carer) exhibit autonomy supportive behaviours (e.g., being empathetic and acknowledging difficulties, giving meaningful explanations, encouraging input, feedback and questions) individuals are more likely to feel a greater sense of autonomy, competence and relatedness, more autonomous motivation and greater behavioural persistence (Ng et al., 2012). A model for long-term engagement based on SDT is visually represented in Figure 2.

**Figure 2.**
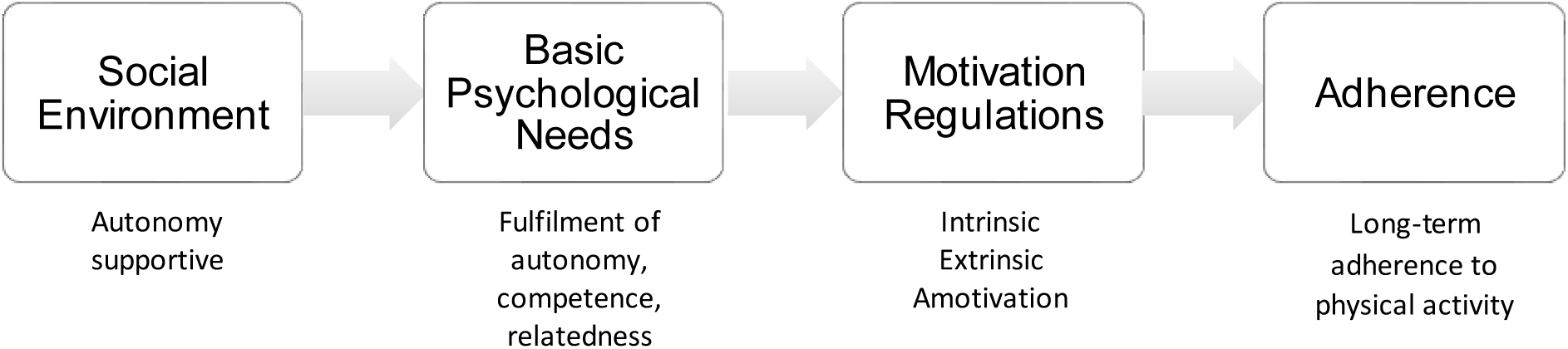
A model of the main propositions of SDT (Ryan & Deci, 2017).

Using SDT as a guiding framework, the present study aimed to explore the motivational process involved in long-term engagement to a home-based exercise and activity intervention Promoting Activity, Independence and Stability in Early Dementia (PrAISED; Harwood et al., 2018) over 24 months. The following SDT propositions were tested:

1. An autonomy supportive context facilitates satisfaction of the three psychological needs;
2. Satisfaction of the basic needs for competence, autonomy and relatedness will facilitate the development of more autonomous motivations (intrinsic, integrated, identified regulations);
3. Individuals with more autonomous motivations will display greater engagement to physical activity and exercise over time, compared to those with more controlled motivations.

## Method

### Design

A longitudinal multiple case study approach was used. A case study methodology is ideal for in-depth exploration of complex phenomenon, and examination of the mechanisms (i.e. answer the how’s and why’s) at play in real-life contexts (Yin, 2018). Further, the multiple case study approach enables exploration of shared patterns of meaning and differences in motivational processes within and between cases and allows for testing of theoretical propositions (Yin, 2018). Ethical approval was granted by the NHS Health Research Authority (Yorkshire and The Humber - Bradford Leeds Research Ethics Committee; reference 16/YH/0040). Written informed consent was gained from all participants prior to involvement. Pseudonyms have been used in this paper to safeguard anonymity.

### Intervention

PrAISED was a person-centred exercise and activity intervention which aimed to increase activity and independence and reduce falls in people with MCI and early dementia (Harwood et al., 2018; Booth et al., 2018). The intervention was delivered by a multi-disciplinary team of physiotherapists, occupational therapists and rehabilitation support workers, at the participant’s home. Participants were encouraged to undertake both tailored strength and balance exercises (referred to as PrAISED exercises) at least 3 times a week and to increase their general physical activity levels via engagement in sport, hobbies and/or general activities of daily living (referred to as physical activities). See Figure 3 for more details. SDT was one of the main psychological theories used to inform intervention development (Booth et al., 2018, 2019). The theory informed the content (with activities designed to satisfy participants’ basic psychological needs, e.g., graded tasks tailored to individuals’ abilities promote competence and person-centred goal setting in which participants chose what physical and/or functional activities they focused on to promote autonomy) and the style of delivery (clinicians received training on need-supportive communication strategies).

**Figure 3.**
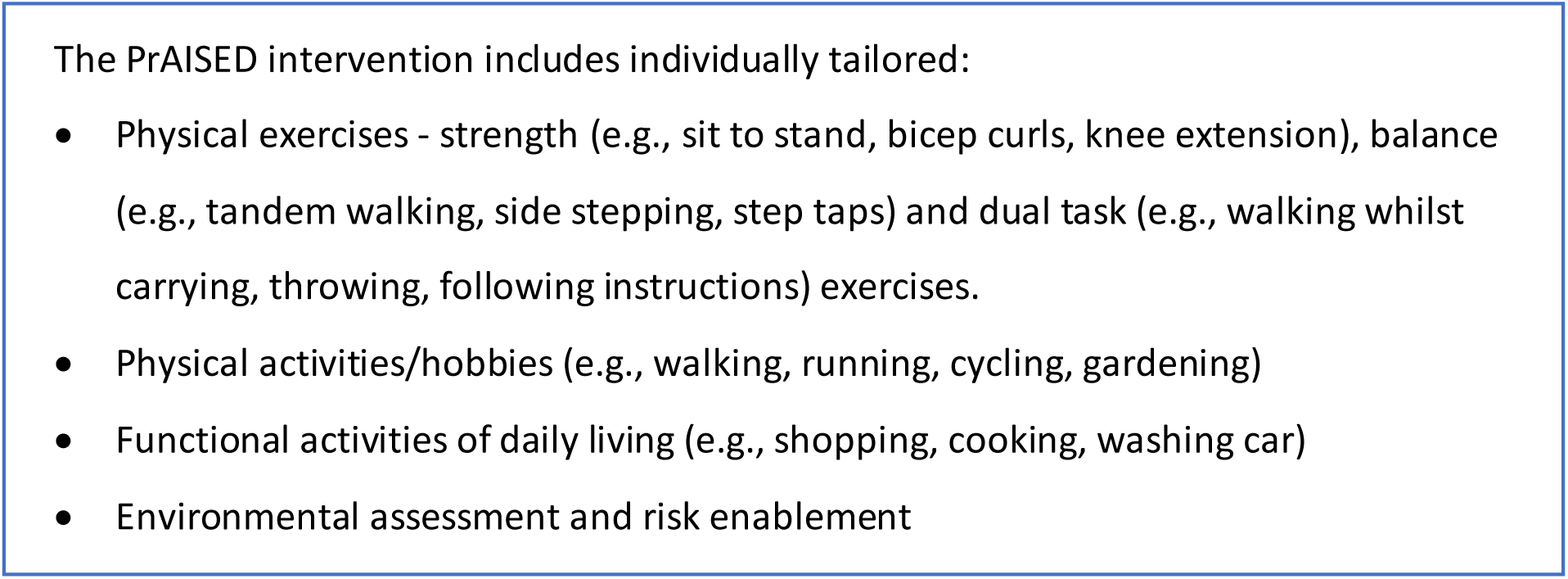
PrAISED intervention

There were two intervention groups, providing moderate (3-month) and high (12-month) levels of supervision for the intervention. It was intended that participants in the 3-month intervention would receive 14 contacts from clinicians (eleven face-to-face contacts plus three telephone calls), with 2 visits in the first week followed by weekly visits thereafter. Participants in the 12-month intervention were planned to have 51 face-to-face contacts, initially delivered twice weekly for 3 months, then tapered to once a month, by month 12. Both groups of participants were encouraged to continue with PrAISED exercises and activities after the intervention had finished.

### Participants

Participants in this study were a subsample of 12 individuals (10 male, 2 female; mean age = 76.6 years, range = 69-91 years) taking part in the PrAISED feasibility trial and their carers (10 female, 1 male). One carer was unavailable to take part in the research due to work commitments. All participants had a diagnosis of MCI or early dementia (made by memory clinic clinicians). For further details regarding recruitment and randomisation please see Harwood et al. (2018). Cases were purposively recruited to obtain a balance of views from those taking part in the 3-month (n = 6) and 12-month interventions (n = 6) and a range of levels of engagement. See Table 1 for participant demographics.

**Table 1.**
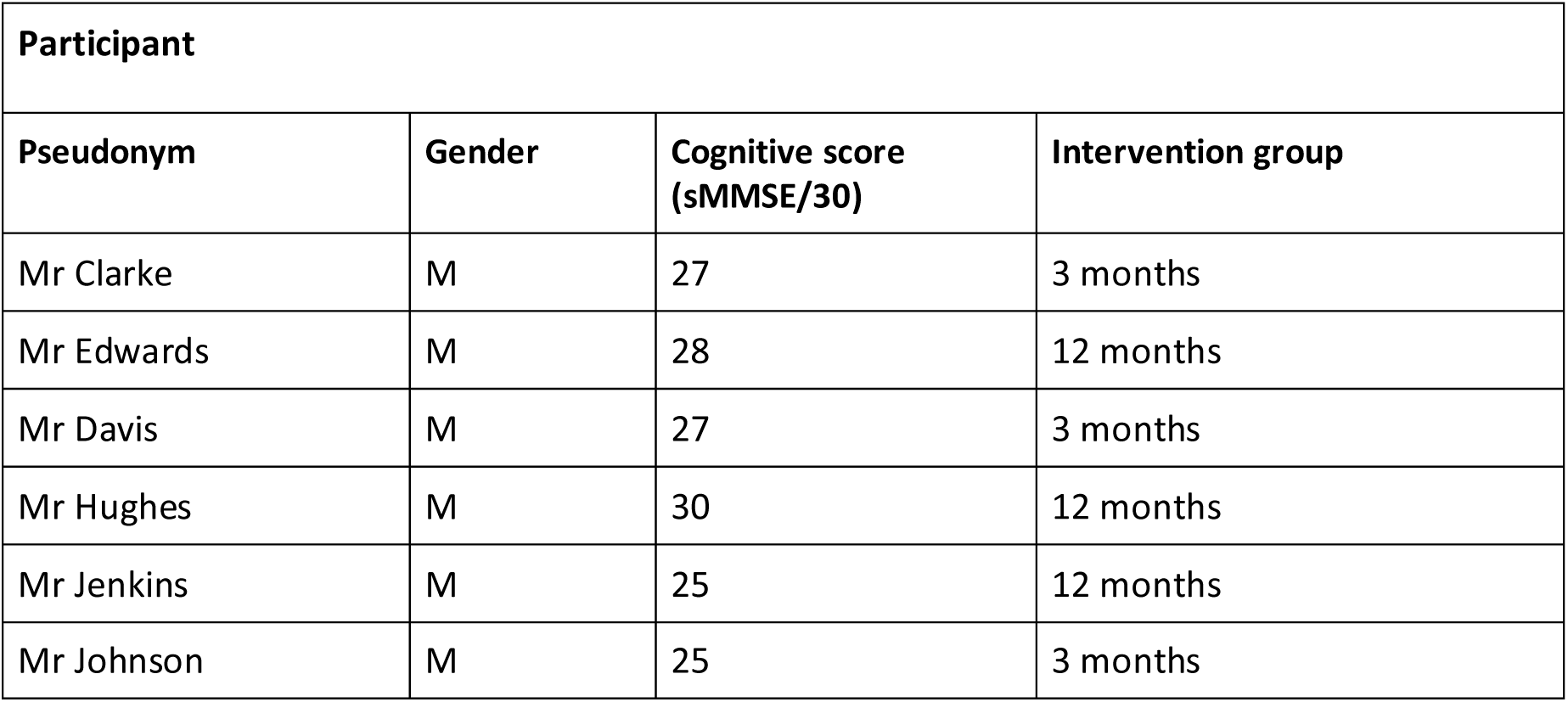
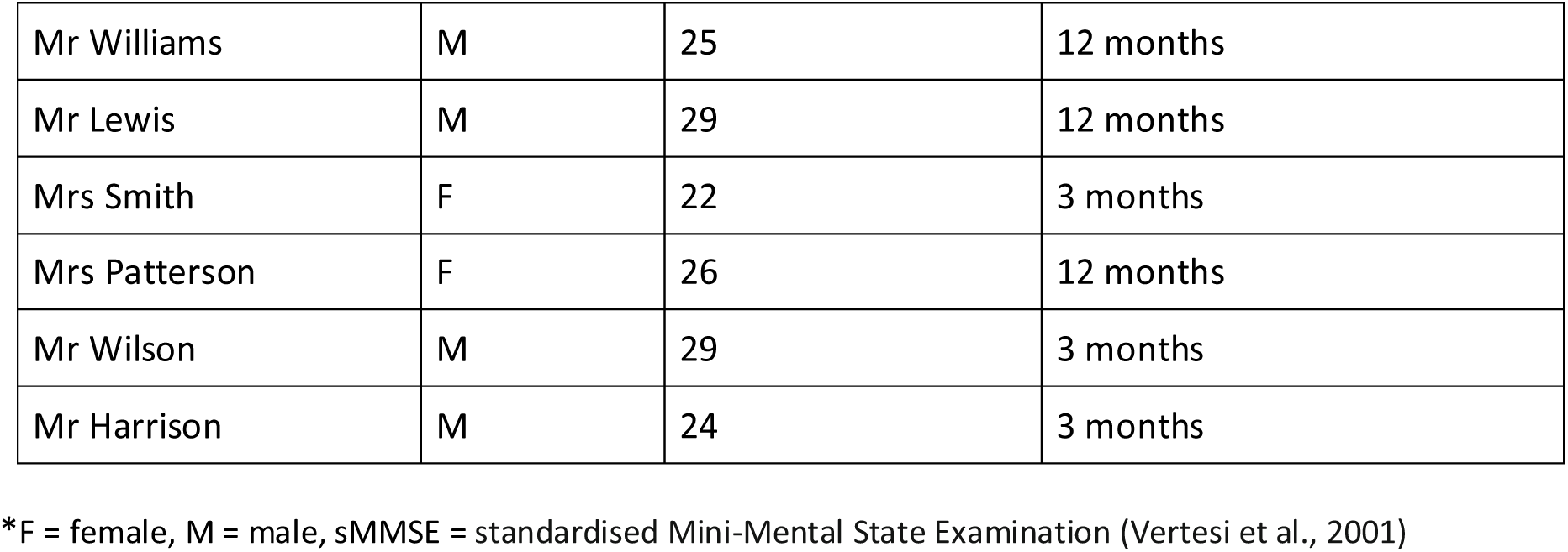
Study participant demographics

| Participant |  |  |  |
| --- | --- | --- | --- |
| Pseudonym | Gender | Cognitive score (sMMSE/30) | Intervention group |
| Mr Clarke | M | 27 | 3 months |
| Mr Edwards | M | 28 | 12 months |
| Mr Davis | M | 27 | 3 months |
| Mr Hughes | M | 30 | 12 months |
| Mr Jenkins | M | 25 | 12 months |
| Mr Johnson | M | 25 | 3 months |
| Mr Williams | M | 25 | 12 months |
| Mr Lewis | M | 29 | 12 months |
| Mrs Smith | F | 22 | 3 months |
| Mrs Patterson | F | 26 | 12 months |
| Mr Wilson | M | 29 | 3 months |
| Mr Harrison | M | 24 | 3 months |

\*F = female, M = male, sMMSE = standardised Mini-Mental State Examination (Vertesi et al., 2001)

### Data Collection

Multiple data sources, collected at different time points over the 24-month study, were used for triangulation and to establish a timeline of events. An evidence database was utilised to enable storage of the data in an organised and accessible form (Yin, 2018). A clear audit trail of analytic decisions was kept to maximise transparency and ensure credibility and quality.

#### Exercise diaries

Participants (with support from their carer where necessary) completed a 12-month diary recording the total number of minutes of PrAISED exercises per day and any other physical activities undertaken (Harwood et al., 2018). Participants returned the diaries to the research team monthly via post.

#### Interviews

The first author (JH) undertook semi-structured face-to-face interviews with participants and their carers in months 4 and 13 in their homes. The participants were given the option to be interviewed individually or together. Eleven dyads were interviewed together, and one participant individually as their carer was unavailable. A semi-structured interview guide was created based on the research teams’ expertise in exercise psychology and feedback from a dementia-specialist Patient and Public Involvement group. Questions aimed to explore participants’ and their carers’ experience with the intervention (e.g., Have you experienced any particular challenges or difficulties in taking part?), motivations (e.g., What are your main reasons for doing the exercises/activities?) and views on the support provided (e.g., Can you tell me a bit about the support you received from the therapists to do the exercises and activities?). The interviews lasted approximately two hours and were audio-recorded. Following each interview, notes were taken of any observations (e.g., non-verbal expressions) that might have been helpful for interpreting the data.

#### Telephone calls

Long-term engagement was assessed by telephone calls at 18 and 24 months. Participants were asked the question: “Have you continued with the PrAISED exercises?” If yes, participants were asked: “How many times a week do you do the exercises?” and “How long (in minutes) do you exercise at each session?” Notes were made detailing any additional information provided by the participant regarding types of activity and reasons for discontinuation of PrAISED exercise or physical activities.

### Data analysis

For the exercise diaries, the number of minutes of PrAISED exercises undertaken per day for each participant were entered into an excel spreadsheet. The following information was used as proxy for adherence:

1. Average number of minutes of PrAISED exercises undertaken per week;
2. Average number of sessions per week;
3. Average duration of sessions;

The mean number of PrAISED exercise sessions undertaken per month was calculated and a graph produced showing each participant’s adherence trajectory overtime. Information on other physical activities undertaken were used qualitatively to inform the case studies.

The qualitative interviews were transcribed verbatim and analysed using a hybrid process of deductive (based on the SDT proposition) and inductive (data-driven) thematic analysis (Braun & Clarke, 2006). A draft summary of each case was written by the first author and reviewed by the clinicians involved in the case.

Data were managed using NVivo software (QSR, 2012), with all sources of evidence organised by case (i.e. participant with dementia). Data analysis was performed using pattern matching logic (Trochim, 1989), in which each case study was compared with the theoretical propositions of SDT. If the case did not fit the propositions of SDT then rival explanations were explored. The framework method was used to facilitate the process (Byrne, 2009). A spreadsheet was generated with columns containing the propositions of SDT and rows of individual cases. Summarised findings from each case were inserted in the appropriate cell. Cross-case synthesis (Yin, 2018) was then undertaken to determine whether any common patterns (i.e. participants’ profiles) around engagement emerged.

## Results

Based on the data, four participant engagement profiles were identified:

1. Long-term engagement - continuation of PrAISED exercises and physical activities for 24 months;
2. Long-term partial engagement - stopped the PrAISED exercises after 12 months, but continued the physical activities for 24 months;
3. Short-term engagement - stopped the PrAISED exercises and physical activities after 12 months;
4. Lack of engagement – PrAISED exercises and physical activities discontinued by 6 months.

The theoretical propositions of SDT were used to guide exploration of the motivational processes involved in participation in the PrAISED intervention and whether these differed based on level of engagement. The findings are presented in light of each SDT proposition with consideration of differences between profiles. A summary is presented in Table 2.

**Table 2.** Summary of each engagement profile and findings in relation to SDT propositions

| Profile | N (%) | Pseudonyms | Findings in relation to SDT theoretical propositions |
| --- | --- | --- | --- |
| 1. Long-term engagement | 2 (17%) | • Mr Clarke<br>• Mr Edwards | • Perceptions of clinicians and carer as need supportive<br>• Needs satisfaction<br>• More autonomous motives for exercises and physical activities |
| 2. Long-term partial engagement | 5 (42%) | • Mr Davis<br>• Mr Hughes<br>• Mr Jenkins<br>• Mr Johnson<br>• Mr Williams | • Perceptions of clinicians as need supportive<br>• Decreased need satisfaction for intervention exercises, increased need satisfaction for other physical activities<br>• More controlled motivations for exercises and autonomous motives for physical activities |
| 3. Short-term engagement | 4 (33%) | • Mr Lewis<br>• Mrs Smith<br>• Mrs Patterson<br>• Mr Wilson | • Perceptions of clinicians as need supportive (but to a lesser extent than profiles 1 & 2)<br>• Lack of need satisfaction<br>• More controlled motives (external and amotivation) for exercises and physical activities |
| 4. Lack of engagement | 1 (8%) | • Mr Harrison | • Low perceived need support from clinicians<br>• Lack of need satisfaction<br>• Amotivation for the exercises |

### Proposition 1 - An autonomy supportive context facilitates satisfaction of the three psychological needs

When asked what made them feel more motivated to do the PrAISED exercises and activities participants, in profiles 1, 2 and 3 (to a lesser extent) reported to perceive a range of need supportive strategies to be exhibited by the clinicians including: taking time to understand the individual and their carer, involving participants and carers in goal setting, actively listening and responding appropriately, acknowledging the participants’ and carers’ feelings, encouraging questions and input, tailoring pace of progression and difficulty level to the individual, providing rationale, giving praise and constructive feedback, and providing support and advice with other health issues. Participants also reported that having similar interests and a shared sense of humour with clinicians to be key motivating factors.

Participants in profiles 1 and 2 tended to report higher levels of need satisfaction than those in profiles 3 and 4 with many reporting need supportive strategies exhibited by the clinicians to facilitate satisfaction of their basic psychological needs.

**Profile 1.** Autonomy supportive strategies were found to aid identification of meaningful goals and thus supporting participants’ feelings of ownership (autonomy). For example, one participant and his spouse explained that they valued the clinicians taking time to understand them, goal setting being a joint process and being given the opportunity to identify meaningful exercises and activities. Continuing to go for walks independently on a daily basis was very important to the participant and something that he enjoyed doing.

However, both he and his spouse were concerned about him getting lost or falling when out on his own. The autonomy supportive strategies helped facilitate a joint discussion that led to the decision that continuing walking independently would be one of the participant’s goals.

> *They [the clinicians] were just really good on picking up on stuff…Because I said I was sort of worried in case he fell while he was out, I wouldn’t know where he was.* [Mr Clarke’s carer, month 4]
>
> *The ones about the routes. That was suggested. Yeah, thought it was a good idea*.
>
> [Mr Clarke, month 4]

The use of autonomy-supportive strategies (e.g., tailoring the pace of progression and difficulty level to the individual) was also important for supporting participants’ basic psychological needs during the intervention. For example, one participant reported feeling a lack of competence for certain exercises, finding some tasks anxiety-provoking. However, the clinicians adapted the exercises to a more manageable level (e.g., reducing repetitions), which the participant felt more comfortable with, thereby enhancing their sense of competence.

Both participants in profile 1 also reported need support from their carers to have been crucial in ensuring continuation of exercises and physical activities.

> *We’ve always supported each other in what effort we do anyway, so, it’s just one of those things.* [Mr Clarke’s carer, month 4]
>
> **Profile 2**. Participants in profile 2 all reported the clinicians to be autonomy supportive. Mr Johnson explained how the need supportive strategies (e.g., using inclusive language, actively listening) contributed to increased feelings of relatedness and autonomy.
>
> *They’ve [the clinicians] got this … attitude, really, that you felt as if you were more or less leading the thing that you were doing. You know, they didn’t say ‘Do this, do that’. It was, well, ‘let’s have a look at this together’, and things like that. And it was, you know, a dual thing that they were there to put ideas forward if you needed them, and just like, somebody coming round for a cup of tea, more or less, you know, a friend.* [Mr Johnson, month 4]

Similarly to profile 1, autonomy support from clinicians aided identification of meaningful physical activity goals. All profile 2 participants identified goals related to re-engagement in an activity (e.g., walking, running) that they had previously enjoyed/valued but for which they were currently not able or motivated to do. Thus, autonomy supportive strategies were particularly important in the earlier stages of the intervention, especially for those who were not keen on prescribed exercises. For example, Mr Hughes initially felt that the PrAISED exercises were boring and he was unwilling to do them (amotivated). It was only when the clinicians used autonomy supportive strategies (e.g., taking time to understand the individual, actively listening and responding appropriately, and encouraging input) and developed a sense of relatedness with Mr Hughes that he explained that all he really wanted was to be able to run again. His carer explained:

> *[the clinician] was obviously listening and said…what would you like to do? And it was the fact that she listened and was empathetic towards the fact that it was no good bullying him to try and do the original exercises but to find something that suited him.* [Mr Hughes’ carer, month 13]
>
> Mr Hughes mentioned how autonomy supportive strategies enhanced his feelings of relatedness with the clinician and helped develop his confidence to run again:
>
> *I’ve a lot of time for [the clinician]. She’s the one who stimulated me. If I were looking for advice and guidance I would turn to her… I find her inspirational…She seems to understand my mental attitude, she understands my physical limitations, she understands what I can do, how to give me positive help. [Mr Hughes, month 4, profile 2]*

**Profile 3.** For participants in profile 3, their motivation for doing the exercises appeared to be mainly driven by the sense of relatedness they felt when with the clinicians. For example, Mrs Smith and her carer reported the clinicians taking time to get to know them and understand their family life:

> *Every time she saw her it was a continuity with the conversations. Got involved with our family lives, and vice versa…talking, chatting with her*. [Mrs Smith’s carer, month 4]
>
> *Oh, it’s a bit daunting, isn’t it, that you’ve got to rely on these people, you know, but never felt it for long, you know, they’re so good. They’re all like one big family*. [Mrs Smith, month 4]

It followed that, although the support from clinicians motivated participants to be active during visits, participants in this profile group often found it challenging to maintain activity levels between visits due to lower levels of autonomy and competence.

> *Mrs Smith responded probably better to the clinicians. They went through the exercises with her every time they visited really, which was a success… she responds easier when there’s someone here to do it*. [Mrs Smith’s carer, month 4].

Other carers in this profile also found it difficult to motivate participants to do the exercises in between clinician visits:

> [carer]: *I’m encouraging [the participant] to do the exercises and, but, I can’t force her to do the exercises*.
>
> [participant]: *Well, she tries, but it’s not the same as [the clinician] coming and doing them with me. I enjoyed [the clinician] visiting. She’s good company…She’s very good at encouraging me. And she makes it more jolly she does it with me, so I do it twice a week with her.* [Mrs Patterson, month 4]

Mrs Patterson explained how she felt confident doing the exercises initially, when she received regular visits from the clinicans. However, there came a point when she no longer felt competent with the PrAISED exercises. This was due to a combination of the exercises being progressed, the clinician visits becoming less frequent (due to the tapered nature of the intervention) and Mrs Patterson’s health continuing to decline.

> *I did them for a little while, and then I dropped them… I could remember the first lot I had, the second lot I can’t remember very much about. I got into a routine at one point, and was doing the first lot quite regularly, in fact, very regularly, every day…It tailed off… I suppose, the others became more difficult.* [Mrs Patterson, month 13]

Participants’ health conditions also greatly limited their willingness and ability to engage in the exercises and physical activities. For example, in addition to a diagnosis of dementia, Mr Wilson also had various other health conditions, and explained that whether or not he did the exercises depended on how he was feeling physically on the day. He reported doing the exercises less in the winter due illness which influenced the extent to which he felt able to do the exercises (decrease in competence).

> *Because it’s colder, you’re getting more kind of illnesses which is making you less likely to want to [do the exercises]…And then when he was ill, well, you had to leave them [the exercises] then*. [Mr Wilson’s carer, month 13]

Thus, provision of need supportive strategies from clinicians were not the only factors that influenced participants’ need satisfaction. At some point during the 12-month programme all four participants in Profile 3 reported a decrease in competence and autonomy in relation to the PrAISED exercises and activities. These feelings appeared to stem from a combination of deterioration in physical health, illness, increased fear of falling, depression and apathy.

**Profile 4.** The participant in profile 4 who disengaged at month 6, did not perceive any specific need-supportive strategies or mention the clinicians to say or do anything that made him feel motivated to do the PrAISED exercises or activities.

> *It was a bit of a waste of time [the clinician] coming, quite honestly… Well, they just virtually went over the exercises and that was it*. [Mr Harrison, month 4]

Mr Harrison exhibited a lack of need satisfaction. He explained that he did not feel that the exercises were optimally challenging and that they did not satisfy his need for competence. He did not report feeling connected to the clinicians or any sense of relatedness.

Furthermore, Mr Harrison mentioned that he did not understand how the exercises would help to improve his memory and resented his partner for ‘nudging’ him to do it. This thwarted his sense of autonomy and negatively impacted his motivation.

> *I’m a man, I guess, and if I’m told to do something, the natural resistance is to say ‘On yer bike’*. [Mr Harrison, month 4]

### Proposition 2 - Satisfaction of the basic needs for competence, autonomy and relatedness will facilitate the development of more autonomous motivations (intrinsic, integrated, identified regulations)

In support of proposition two, participants who reported increasing levels of need satisfaction (profiles 1 & 2) also reported increases in more autonomous motivation for the PrAISED exercises and/or activities. Furthermore, individuals with lower levels of need satisfaction or who experienced a decrease in need satisfaction tended to report increased controlled motives.

**Profile 1**. Both participants in profile 1 over time described increasing feelings of confidence doing the exercises and activities (competence - “*Challenging, but yet at the same time, I do find a bit of enjoyment from it [the PrAISED exercises]…There’s confidence in doing it”,* Mr Edwards), ownership (autonomy – *“I’ve written all these on [notes on how to do the PrAISED exercises]…I switched the sequence, rather than, you know, have weights on and have to take them off, and then put them back on again afterwards, I put all the weight ones together”, Mr Clarke*) and connection (relatedness - “*You tend to have nice chats with other walkers, it’s a good way of socialising*”, Mr Edwards’ carer). Participants in profile 1 also reported an increase in autonomous motives and decrease in controlled motives for the PrAISED exercises and activities over time. For example, in the initial stages of the intervention both participants reported a mix of autonomous and controlled motives for engagement. They wanted exercises and activities that could help them cope with a diagnosis of dementia, overcome depression and delay cognitive decline (identified regulation) but also felt an obligation to themselves and the research team (introjected & external regulation). However, when interviewed in month 13, both participants’ motivations had become more internalised. For example, Mr Clarke no longer felt externally motivated and instead reported doing the exercises on a daily basis, mainly because he enjoyed it.

> *I think initially it was, because you had to fill it [the daily exercise diary] in. But, I think it’s become such a routine now.* [Mr Clarke’s carer, month 13]
>
> *Well, I enjoy it, and it’s, I think its good health wise. It’s become a way of life now… I do them religiously every day.* [Mr Clarke, month 13]

**Profile 2.** In support of proposition 2, satisfaction of participants’ needs for autonomy, competence and/or relatedness seemed to enhance participants’ autonomous motives for doing the exercises. For example, Mr Hughes, as previously described, started the programme amotivated for doing the PrAISED exercises. However, satisfaction of his needs for relatedness and autonomy through joint identification of a meaningful goal (i.e., running) led him to identify with the benefits of doing the exercises and thus his motives became more autonomous.

> *They’ve asked me now if I’d like to have additional exercises. I’ve said yeah, if it helps me to straighten that hip, let’s do it. Because the running now it becomes quite an important part… I feel I’m preparing myself to run tomorrow*. [Mr Hughes, month 4]

All participants in profile 2 reported a drop in at least one of the basic needs for the PrAISED exercises at some point during the 12-month period. Two of the participants had a trip or fall, and one had minor stroke, which led them to feel less confident doing the exercises (reduction in competence), four of the participants no longer saw the purpose of doing the exercises (decrease in autonomy) once their injury had healed/they had increased in mobility enough to do the activities that they enjoyed doing, and all reported missing the clinicians doing the exercises with them (drop in relatedness). Thus, a decrease in need satisfaction resulted in lower levels of self-determined engagement for doing the exercises.

Satisfaction of basic needs also appeared to enhance or sustain participants’ feelings of intrinsic motivation for engaging in meaningful physical activities identified as part of the PrAISED programme. For example, one participant described enjoying daily walks with his spouse to the local shop as it gave him a sense of purpose (autonomy satisfaction) and opportunities for social connection (relatedness satisfaction). Following a physical injury, this participant reported a temporary lack of confidence in their ability to carry out the activity (lack of competence). However, engaging in the programme’s exercises helped restore physical confidence (competence satisfaction), enabling continued participation in an activity that was both enjoyable and personally meaningful.

**Profile 3.** Similarly to those in profile 2, satisfaction of participants’ basic needs did appear at times to lead to more autonomous motives. For example, two participants in profile 3 described the PrAISED exercises and physical activity as occasionally enjoyable (intrinsic motivation), given the social interaction (relatedness) with clinicians.

[carer]: *…and there are times when you say, you’ve enjoyed it*.

[participant]: *Yeah. Nearly always. Just refreshed me really*.

Interviewer: *What do you enjoy about it?*

[participant]: *Well, just, everybody’s laughing all the time.* [Mrs Smith, month 4]

However, as previously described over time all four participants in Profile 3 reported decreasing feelings of competence and autonomy in relation to the exercises and physical activities. Deterioration in health meant that carer support was essential for these participants to maintain engagement over time. However, the participants often felt their autonomy to be thwarted when family carers encouraged them to exercise, which created tension in the relationship and an increase in more controlled motives (external regulation).

**Profile 4.**

In line with SDT, Mr Harrison described low levels of need satisfaction and thus his motives for engagement were more controlled. Mr Harrison expressed a strong dislike for the prescribed exercises, believing them to be boring. He stated that he preferred other activities and indicated that he was extrinsically motivated, only complying with the regime not to let the research team down.

### Proposition 3 - Individuals with more autonomous motivations will display greater engagement to physical activity over time, compared to those with extrinsic motivation

Proposition three was supported as participants reporting more autonomous motives for the PrAISED exercises and/or activities (profiles 1 and 2) were more likely to continue long-term (for more than 12 months) compared to those in profiles 3 and 4 who reported more controlled motives for engagement. Participants in profiles 3 and 4 only engaged in the PrAISED exercises and activities in the short-term (12 months or less).

**Profile 1.** At 24 months following baseline only two participants reported to have continued to perform the individually-tailored set PrAISED strength and balance exercises. At 13 months both these participants reported still doing the exercises on a regular basis because they were enjoyable (intrinsic motivation) and had become an important part of their routine serving some higher purpose/value (e.g., a sense of achievement and aiding management of depression). One participant also mentioned that at times he was motivated by more controlled reasons, feelings of guilt due to concern about letting himself and the research project down (introjected and external regulation). However, he explained that his main motivations for doing the exercises are more autonomous which may be why he reported still doing the exercises 2 times a week in months 18 and 24.

> *If I stopped, then I’ll start criticising myself over it. And say you’re letting yourself down as well as letting down the work of these people that are attempting to help me out… I’m happy to do them. To me it’s a positive thing, it’s an achievement. There’s some benefit coming out if it, it’s getting the brain working, it’s not coming too static.* [Mr Edwards, month 13]

Both participants in this group had also continued engaging long-term in physical activities introduced through the exercise programme, for which they felt intrinsically motivated.

**Profile 2.** The combination of motivation regulations underpinning engagement varied over time for each participant. Participants in profile 2 revealed a mix of autonomous (mainly identified) and controlled motives (introjected and/or external) and even amotivation at times for doing the PrAISED exercises. However, on all occasions when participants described an increase in autonomous motivation this was reflected in their increasing engagement. For example, as Mr Hughes came to view the PrAISED exercises as beneficial for supporting his running, there was an increase in his reported exercise engagement (from 3 times per week in month four to 5 times per week by month six). However, once the clinician visits stopped his engagement reduced to doing the PrAISED exercises on average once a week and it became apparent how important the external support from the clinicians was to motivate him to do the exercises regularly:

> *I just need something to give me that push. I could get up now and do them but I don’t.* [Mr Hughes, month 13]

Towards the end of the programme, Mr Hughes reported losing interest for particular exercises and expressing not seeing the point in doing them (amotivation).

> [participant]: *Rolling around on the floor, well, all right, you know [laughter] bit pointless, waving legs in the air…*
>
> [carer]: *There was a point to all of it. Susan [the physiotherapist] didn’t give you anything that didn’t have a point*.
>
> [participant]: *Oh aye, yeah, I understand that, yeah. But, there may have been a point to it but I, maybe I didn’t see the point.* [Mr Hughes, month 13]

Mr Hughes’ case highlights the dynamic nature of motivation regulations and how they can fluctuate overtime. For all participants in profile 2, once they had successfully re-engaged in their chosen activities (e.g., running, walking, and gardening) they no longer viewed the PrAISED exercises as of value (reduction in identified regulation). Thus, the PrAISED exercises were seen as facilitators to doing the activities that they enjoyed doing rather than as something that is of benefit for its own sake and thus were not continued longer than 12 months.

Motivation for the physical activities undertaken as part of the PrAISED programme (e.g., walking, running, gardening) were generally more autonomous for those in both profiles 1 and 2 and hence were continued long-term (greater than 12 months).

> *I’m not doing it [the PrAISED exercises] in the house, I go walkies. I’ve been lucky that I’ve got neighbours that have got dogs who like to get out virtually every other day. And we done a few miles with them. I enjoy doing it…Not only that, you can do something in that walking, like the shopping, like the gardening, like dog-walking. I’m achieving something, as well as achieving myself*. [Mr Jenkins, month 13]

**Profile 3.** Participants in profile 3 reported mainly external motives for the exercises and physical activities.

> *If my wife didn’t come home and get me to do it, I probably wouldn’t.* [Mr Lewis, month 4]

Similarly to those in profile 2, engagement stopped as soon as the external motivators (support from clinicians and regular checks from the research team) ceased.

The participants also reported frequent health problems, which in three cases deteriorated to such extent that the participants moved to a care home and withdrew from the intervention.

**Profile 4.** In support of SDT, once the clinician visits stopped, Mr Harrison expressed not seeing the point in doing the PrAISED exercises (amotivation) and reported having discontinued the exercises, on the grounds that he was already very active and did not want to make any changes to his lifestyle.

He proved however, intrinsically motivated to do the physical activities that he was already taking part in prior to starting the PrAISED intervention. He explained that he highly valued being active in general, enjoyed it, and believed keeping moving to be key to maintaining physical and mental health and well-being.

## Discussion

A longitudinal multiple case study was conducted to explore the motivational processes involved in long-term engagement to a home-based exercise and physical activity intervention (PrAISED). The main objective was to test the applicability of the SDT propositions with older adults with MCI and early dementia.

### Proposition 1

This study found support for proposition 1 that autonomy supportive strategies from clinicians contributed towards satisfying participants’ feelings of basic need satisfaction. In the context of this study, need support from clinicians early on in the intervention was critical to engagement. Previous research testing a walking intervention in adults (Kinnafick et al., 2014), reported satisfaction of the needs for competence and relatedness to be critical in the adoption stage, with autonomy becoming more important in the adherence phase. In contrast, the current study found support for participants’ autonomy, through identification of meaningful activities, was critical early on the intervention. It may be that the greater functional significance of autonomy in the adoption phase of the intervention was due to the tailored nature of the intervention. In the study by Kinnafick et al. (2014), participants signed up to a walking intervention and the type of physical activity was set. However, in the PrAISED intervention the types of exercise and activities undertaken were tailored to the individuals’ interests and needs.

It is important to note that factors other than clinician autonomy support (e.g., illness, increased fear of falling, depression and apathy) also appeared to influence the extent to which participants felt autonomous and competent at performing the PrAISED exercises and activities unsupervised between visits. Thus, future studies with this population should consider monitoring these factors and exploring further how to most effectively support individuals experiencing these barriers. For instance, fear of falling appeared to become more of an issue when the exercises were progressed beyond what the individual felt comfortable doing unsupervised. Although it is advised that exercises are progressed (in intensity or challenge) in order to have beneficial effects on balance or mobility (Sherrington et al., 2017), individuals with a high fear of falling may be more likely to continue with the exercises long-term if the exercises are kept at a level the individual feels comfortable with. Even if the difficulty is not enough to result in improvements in balance or mobility, some exercise is considered to be better than no exercise (Piercy et al., 2018) as it may provide other health benefits (e.g., improved mood, quality of life, and blood pressure).

Furthermore, there were individual differences in the extent to which depression was a barrier to activity of not. Interestingly, depression and apathy were barriers for participants in Profile 3 but not for those in Profile 1 who used the PrAISED exercises as a way of managing their depression. It may be that helping individuals to re-frame exercise and/or activity as a way of managing depressive symptoms and employing behavioural change techniques that target emotion (Glowacki et al., 2017) may be beneficial.

### Proposition 2

In support of proposition 2, satisfaction of participants’ basic needs seemed to enhance participants’ autonomous motivation for doing the PrAISED exercises and activities. The longitudinal nature of the study enabled examination of fluctuations in participants’ basic need satisfaction and motivation regulations over time. Similarly to previous longitudinal research (Rahman et al., 2011), change in need satisfaction appeared to be related to change in motivation regulations.

According to SDT (Ryan & Deci, 2017), humans are naturally inclined to internalise extrinsic motivations and become more intrinsically motivated over time. Studies with older people with normal cognitive function have confirmed this proposition (Lee et al., 2016). However, thus far, it was unclear whether symptoms of dementia (e.g., memory problems and neuropsychiatric symptoms) which influence older adults’ capability and motivation to engage in regular exercise (van Alphen et al., 2016), might forestall the process of internalisation. To our knowledge, this is the first study testing the SDT proposition in a population with MCI and early dementia. Participants in profiles 1 and 2 demonstrated that it was possible for individuals in this population to become more autonomous in their motivation for exercises and/or physical activities overtime despite experiencing difficulties dealing with a diagnosis of dementia, depression and cognitive decline. Thus, these findings suggest that SDT is a suitable framework to explore the mechanisms underlying activity persistence in older adults with MCI or early dementia.

A novel finding was that the heavily supervised nature of the research study (i.e., regular clinician visits and exercise diaries), and a strong connection with clinicians (relatedness), appeared to strengthen controlled motivations (introjected and external regulation) for some participants (particularly those in profiles 2 and 3). Although this sense of obligation to the research may have facilitated engagement in the intervention initially, when the exercise diaries and clinicians visits ceased the participants’ main reasons for doing the exercises were no longer there – resulting in a decrease in relatedness, increase in amotivaton and dropout. Kinnafick et al. (2014) also found participants to report a sense of obligation to the walking programme which facilitated physical activity adoption. However, this became less relevant in the later stages of the programme as participants’ motivation became more internalised. Future studies could investigate how involvement in research and heavily supervised interventions may increase participants’ extrinsic motivations for engagement in physical activity and the extent to which the research itself is an intervention. Furthermore, consideration should be given to the implications of rolling out such interventions in the ‘real world’ without the added research supervision and possible implications on individuals’ engagement.

### Proposition 3

In support of proposition 3, only individuals who were more autonomously motivated (Profiles 1 and 2) continued with the exercises and/or activities unsupervised in the long-term. These results were aligned with previous SDT research in other populations which have found individuals with more self-determined motivation to be more likely to engage in desired behaviours long-term (Ng et al., 2012; Rogers et al., 2012; Sarrazin, 2002; Teixeira et al., 2012).

The findings suggest that for an older adult with MCI or dementia to engage in exercise or physical activity long-term the behaviour needs to be enjoyable (intrinsic motivation) and/or serving some other purpose (identified regulation) such as helping cope with depression, or as a facilitator to re-engaging in enjoyable activities. Individuals can possess levels of controlled forms of motivation and persist with an activity (for example Mr Edwards), as long as overall motivation is more autonomous (Markland and Ingledew, 2007).

Individuals with more controlled motives and/or amotivation (Profiles 3 and 4) engaged in the PrAISED exercises and activities in the short-term but discontinued as soon as support (clinician visits and/or exercise diary) came to an end. Thus, findings are aligned with previous research which has suggested that more controlled forms of regulation and amotivation are unlikely to be conducive to long-term activity engagement (Teixeira et al., 2012; Sarrazin, 2002). These findings highlight the importance of consideration of participants’ motivation regulations with individuals ideally being autonomously motivated before support for exercise or physical activity is withdrawn.

### Strengths and Limitations

A novel aspect was the use of a multiple case study methodology to capture the complexity of the phenomenon of exercise and physical activity engagement in older adults with dementia in a real-world context (Yin, 2018, p.270). The use of multiple case study design also enabled testing for theoretical replication and consideration of rival explanations (Yin, 2018, p.177). A limitation of the study was that measures of engagement were based on self-reported data, so findings may have been subject to inaccurate reporting or social desirability bias. In order to counterbalance this risk and in line with good practice in case study research, we used multiple data sources. We are confident that this strategy enhanced data credibility (Yin, 2018) and ensured a fuller range of perspectives be captured in describing the phenomenon.

### Practical and Research Implications

This work presents implications for future research. Future research could test the consistency of the identified profiles through further empirical studies. If generalisability of the profiles is confirmed they could be used to inform tailoring of future interventions and/or aid prediction of adherence risk. However, the dynamic nature of participants’ motivation and the influence of factors outside of the intervention’s control, such as illness or life events, should be taken into consideration. It may be that over time individuals switch between profiles, thus regular monitoring and flexibility in intervention delivery could be necessary.

In terms of implications for clinical practice, findings from this study can be used by researchers, practitioners and policy makers to inform the design and development of future interventions supporting older adults with MCI or dementia to engage in physical activity and/or exercise long-term. In particular, findings highlight the importance of interventions considering not just what is delivered (the content) but how (the communication style used by clinicians). Thus, clinicians should aim to exhibit an autonomy supportive style with the aim of optimising need satisfaction throughout the intervention.

This study has highlighted that a high level of ongoing practical and emotional support and a feeling of connection with clinicians can lead to long-term engagement in exercise and/or physical activity in individuals in with MCI or early dementia. To support engagement future practice could give a person with dementia the right to choose a clinician with which they have a good match (i.e., similar interests, values, and/or humour) and request to change to a different therapist if they feel that sense of relatedness is missing. However, the increasing population size and limited resources means that the feasibility of being able to deliver long-term individualised support to the wider population may be questionable. Thus, there may be value in ‘screening’ for individuals who are most likely to benefit from interventions such as PrAISED. For instance, individuals who are already highly active and intrinsically motivated to engage in regular physical activity (e.g., Mr Harrison, profile 4) are unlikely to need or benefit from an intervention, such as PrAISED. Furthermore, individuals with health conditions and a level of cognitive impairment which greatly limits their ability to engage in exercise and physical activity (e.g., those in profile 3) would likely not benefit without substantial carer support.

Finally, the findings suggest that SDT could be used to understand the motivational processes of engagement to a home-based activity intervention in older adults with MCI and early dementia. However, there are alternative theories to SDT, which could be used to investigate engagement to physical activity in this population. These theories explain behaviour change and maintenance by emphasising other variables or constructs, such as self-efficacy, expectations, or capability. These constructs could be integrated with SDT tenets to develop and field-test a more holistic framework to understand long-term engagement.

## Conclusion

This study provides new insights into the factors influencing long-term engagement of older adults with MCI and early dementia in exercise and physical activity. Aligned with SDT, the provision of basic needs support over time and intrinsic motivation proved to be key factors influencing participants’ long-term engagement to physical activity. Dementia-related factors (e.g., memory problems, multi-morbidity, depression and/or apathy) were found to have a negative influence on engagement for some individuals. These findings can be used by researchers and health professionals to guide future research and practice.

## Data Availability

All data produced in the present study are available upon reasonable request to the authors

## Acknowledgements

We would like to thank the Dementia Patient and Public Involvement group at the University of Nottingham for their input and advice on the interview guide. We would also like to thank Rupinder Bajwa and Dr Sarah Goldberg for their support and the PrAISED participants who generously gave their time to the study.

